# Mining Twitter Data on COVID-19 for Sentiment analysis and frequent patterns Discovery

**DOI:** 10.1101/2020.05.08.20090464

**Authors:** Habiba Drias, Yassine Drias

## Abstract

A study with a societal objective was carried out on people exchanging on social networks and more particularly on Twitter to observe their feelings on the COVID-19. A dataset of more than 600,000 tweets with hashtags like #COVID and #coronavirus posted between February 27, 2020 and March 25, 2020 was built. An exploratory treatment of the number of tweets posted by country, by language and other parameters revealed an overview of the apprehension of the pandemic around the world. A sentiment analysis was elaborated on the basis of the tweets posted in English because these constitute the great majority (USA, GB, India…). On the other hand, the FP-Growth algorithm was adapted to the tweets in order to discover the most frequent patterns and its derived association rules, in order to highlight the tweeters insights relatively to COVID-19.

## 1 Introduction

Coronavirus disease 2019 (COVID-19) was first detected in Wuhan, China, in December 2019 and has spread worldwide in more than 198 countries. Within a couple of months, this outbreak has upset the world and uncertainties about the future were for the first time evoked. Several epidemic periods have been observed in the world. In the recent years, this epidemics phenomenon has grown because of the contagion favored by the globalization. Nowadays, epidemics provoke extreme economic crisis at the scale of countries as well at the individual level. People can reach psychosis symptoms because of the contagion and countries may suffer from economic crisis due to people traveling restriction and social isolation. In [8], the authors give several battle hints to fight the virus as it travels the world, the metaphor of war is used.

Doctors and people the most affected psychologically by the epidemics are the most likely to talk about it on social networks like Twitter, which have become essential in our daily lives. The analysis of internet users’ behavior has shown that they are mostly either seekers of information from the Web or social media (Facebook, Twitter, microblogs…) [3], [4], [15]. Understanding better this data flow can be of great benefit to people and the industry to make informed and focused decisions on how to meet goals. Many companies have monitoring activities on social media sites to collect information. For example, talking on social sites about a city or an airport in a positive way improves its image.

One of the most popular social networks on the web is Twitter. Tweets are based on the format of micro-blogs and constitute an important source of archive data, allowing its export for academic research purposes.

As part of this work, we performed a datamining study on COVID-19 by analyzing Twitter publications in relation to this disease, posted between 23 February 2020 and 03 March 2020. The outcomes could help dressing an inventory for the history of the outbreak and especially how it was apprehended by the world population. Technically, we first collected tweets in relation to the coronavirus using NodeXL and stored the recovered tweets in a dataset. In a second step, the data underwent a preprocessing with a statistical analysis in order to extract knowledge helping their understanding. Thereafter we conducted a sentiment analysis on the dataset to come up with the feelings of the twitters during the period of study. In addition to this effort, the most frequent patterns were extracted with the aim to grasp social features about the twitters. Among the datamining algorithms that can handle such issue, FP-Growth was selected because of its effectiveness and efficiency.

The paper is organized as follows. In the next section, we present related and recent works on tweets analyses and the coronavirus. Section 3 details the approach of building the tweets dataset. Section 4 describes the the sentiment analysis algorithm we designed as well as its empirical results. Section 5 presents the adaptation of FP-Growth algorithm to tweets and the experimental results prior to conclude the main contributions.

## 2. Related Work

COVID-19 appeared recently in December 2019 in Wuhan China and yet hundreds of publications on this topic updated daily can be found in [16]. Studies from around the world are performed within a very short period of time as the COVID-19 took its toll in almost all countries [14]. This fact translates the importance of studying this virus in order to speed up the mastery of the disease and the discovery of a remedy. With an impressive number, popular articles on COVID-19 are widely published on daily classic media relating the disease spread, the worrying issues and also helping to cope with the cultural customs and to urge for people isolation to limit the disease propagation. This invisible virus is considered as the most dangerous enemy for the humanity and the metaphor of war against it, was adopted by certain scientists who advice some military strategies to combat this disease [8].

Tweets analysis has known an increasing number of efforts these last years. Most of them are interested in determining features of social interactions of tweeters [2] [4] and tweeters behaviors [3] [15]. Also tweets analytics tools have been developed to be applied on numerous subjects [17].

There is a recent and rich literature on sentiment analysis (SA) and its applications to various fields. SA has been also investigated for social networking data and is generally used by companies for analyzing the opinion and feelings of the customers about products, services and company strategies [5] [9] [10] [11] [12].

Sophisticated datamining tools such as classification, clustering and association rules mining were developed [7] [13] and used in a very large spectrum of domains. Association rules mining that are investigated on our tweets data have also known impressive development for use on huge volume and complex data [1] [6].

For the present work, as COVID-19 is the subject of the hour for the whole planet, we conducted a tweets analytics study on this phenomenon focusing on generating social features of tweeters hoping to achieve important insights.

## 3. Tweets Collect and building a dataset

As a starting step to handle the study on COVID-19 tweets mining, we carried out the construction of a dataset. We provide metadata for the dataset, which merely describes its characteristics such as its title and description, keywords or tags, publisher, date of publication or terms of use. In this section, we present the different steps performed to achieve a dataset of tweets on COVID-19 of a high quality.

### 3.1. Extraction of raw data

*NodeXL* was exploited to extract tweets related to the Coronavirus topic and highlighted by the following hashtags: #COVID-19, #COVID19, #COVID, #Coronavirus and #Corona. The crawling data period started on the 27^th^ of February and ended on the 25^th^ of March, which is spread over four weeks. During this phase, the world has known tremendous changes, especially in its whole organization. The huge number of infected and dead people provoked public panic and fear, which raised supply shortages not only in pharmaceuticals and PPE but also in food. Several countries have known a bad public health management and a rapid scalability in world economic crisis has been observed during a very short lapse of time. Rushes on public market have been seen before certain governments decreed isolation and quarantine for inhabitants an especially for returning travelers. All these upheavals impact on the humanity behaviors.

The aim of this study is to shed light on all these aspects through mining tasks on the extracted collection. The latter contains more than a half million tweets written in several languages and sent from many countries.

### 3.2 Tweets Preprocessing

Programming codes in Java language were developed for basic cleaning steps on the collected tweets. Textual data analysis was first held to eliminate hyperlinks, mentions and punctuations. Stop words such as the articles, the prepositions of conjunction, time and place and the superfluous and unnecessary words, were removed as they do not have impact on the text meaning. The keywords are then determined by the Stemming Algorithm, which consists in associating to each remaining word its root. Let remark that some words were misspelled, an example is the word ‘carona’ which was used by President Trump and repeated by others users 8013 times. This kind of typos was simply eliminated.

### 3.3 Descriptive Dataset and Metadata

The created dataset includes at the row level the different tweets and at the column level the different attributes, which are described as follows.

1. Author
2. Recipient
3. Tweet
4. Hashtags
5. Language
6. Relationship
7. Location
8. Date
9. Source

The Attributes are all of string type except for the date which follows the ‘month/day/year’ format. The dataset contains 653 996 tweets with no missing values. Table 1 gives an overview of a portion of the dataset with 9 attributes, the contents of the tweets are truncated for lack of space to insert the figure. Note that the location is not always provided.

**Table 1.**
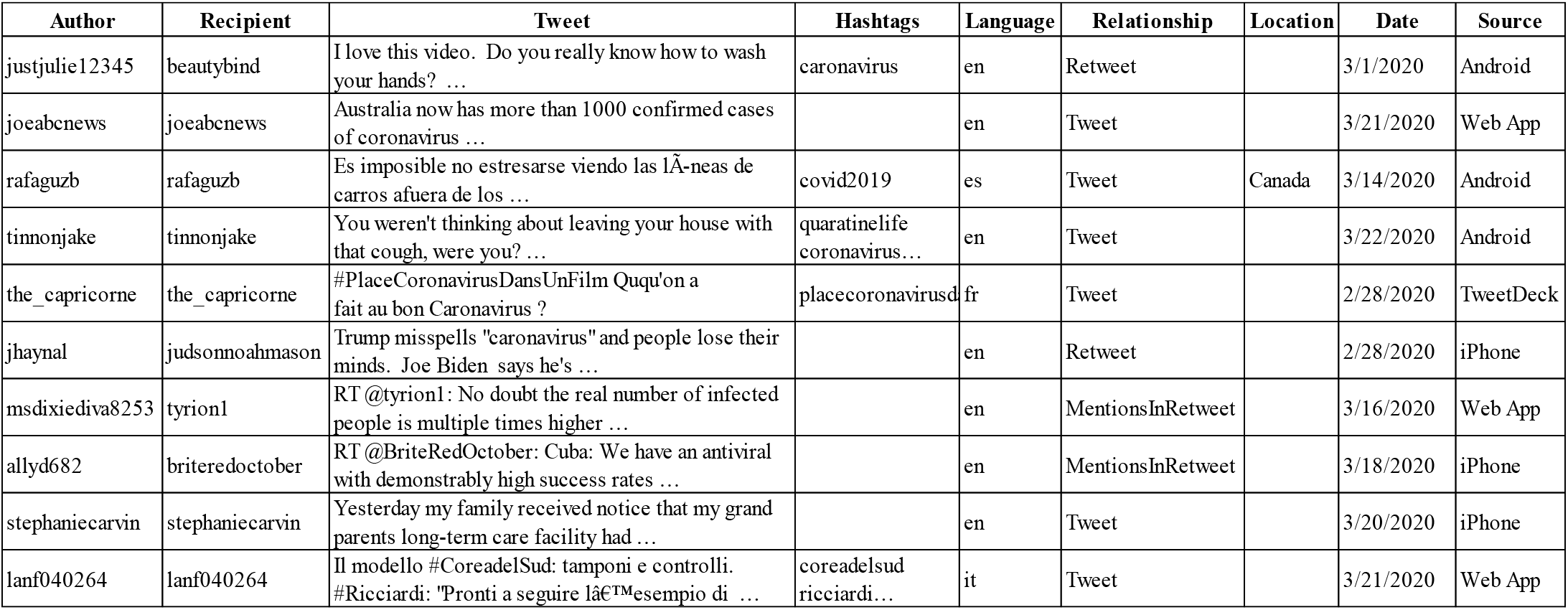
Illustration of the dataset structure.

## 4. Tweets Analytical Insight for COVID-19

As a second contribution, we performed an analytical study on the constructed dataset in order to mine knowledge allowing an understanding and a mastery of the tweeters insights before performing datamining tasks, which help to provide a more comprehensive perception. This analysis provides data trends and distributions, descriptive statistics, data groupings and helps formulating hypotheses. This important and costly step is common to several data mining tasks. Among the latter, we focus on association rule generation based on frequent patterns mining using an adapted FP-Growth algorithm [7].

The experiments of the developed program in Java for analytics was executed on the 653 996 tweets and the different outcomes are exhibited in the following subsections.

### 4.1. Top Hashtags

The hashtags #COVID-19, #COVID19, #COVID, #Coronavirus and #Corona that served for the construction of the dataset are not considered since each tweet contains at least one of them. Figure 1 shows the top 20 hashtags, which report some events and situations such as *pandemic, update, outbreak, stayAtHome, curfew, ConfinementTotal* (French words) and *quarantine*, people are dealing with. On the other hand, countries and regions such as China, Iran, Wuhan, India and Italy, that were the most affected by the virus are cited. Note that although India is knowing the phenomenon at the end of the studied period, we assume that it is evoked among these countries because of its large population.

**Figure 1.**
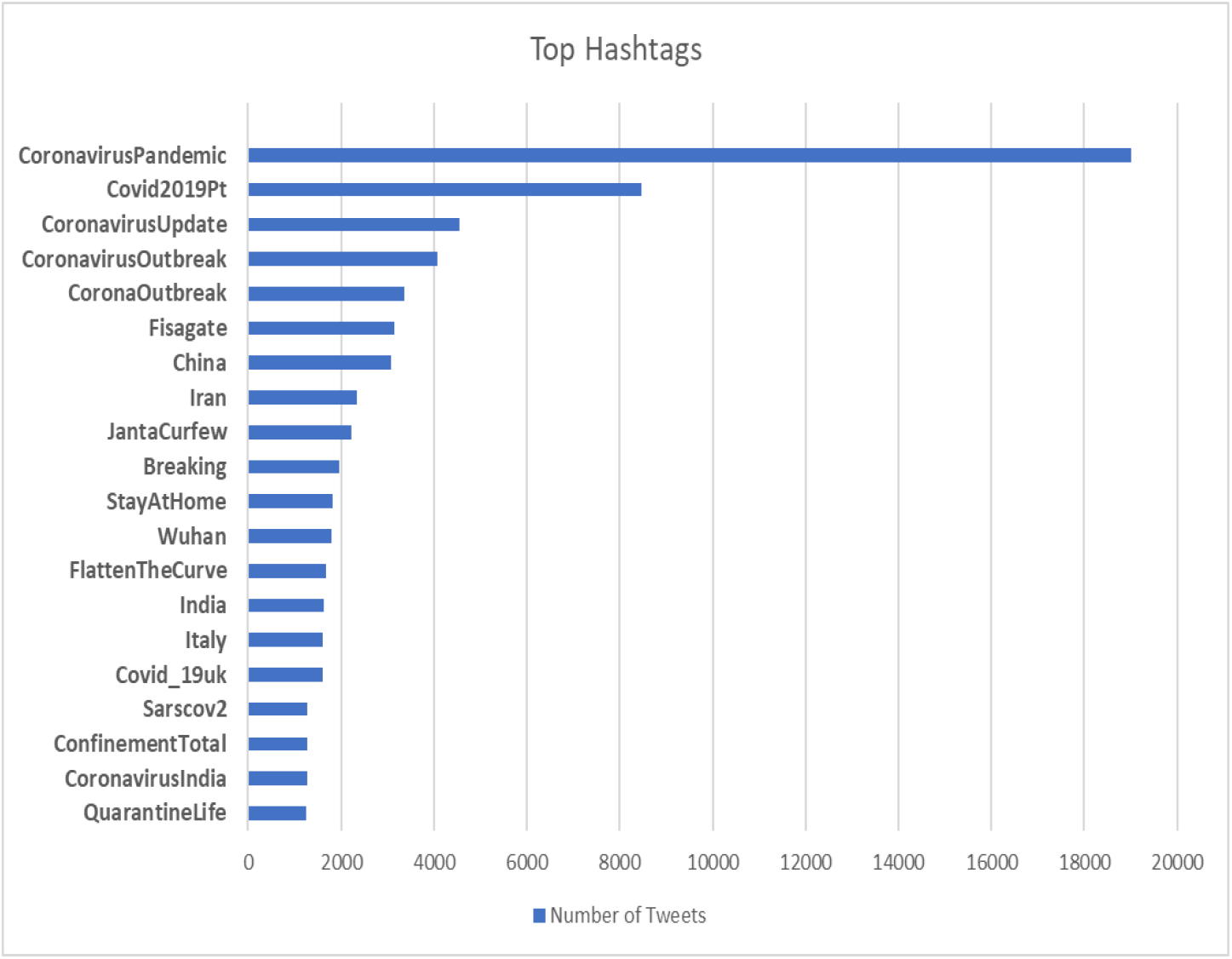
The 20 Top Hashtags of the dataset.

### 4.2. Tweets sources

The sources from which the extracted tweets were published are shown in Figure 2. The most used platforms are Android, iPhone and Web App with approximatively the same ratio.

**Figure 2.**
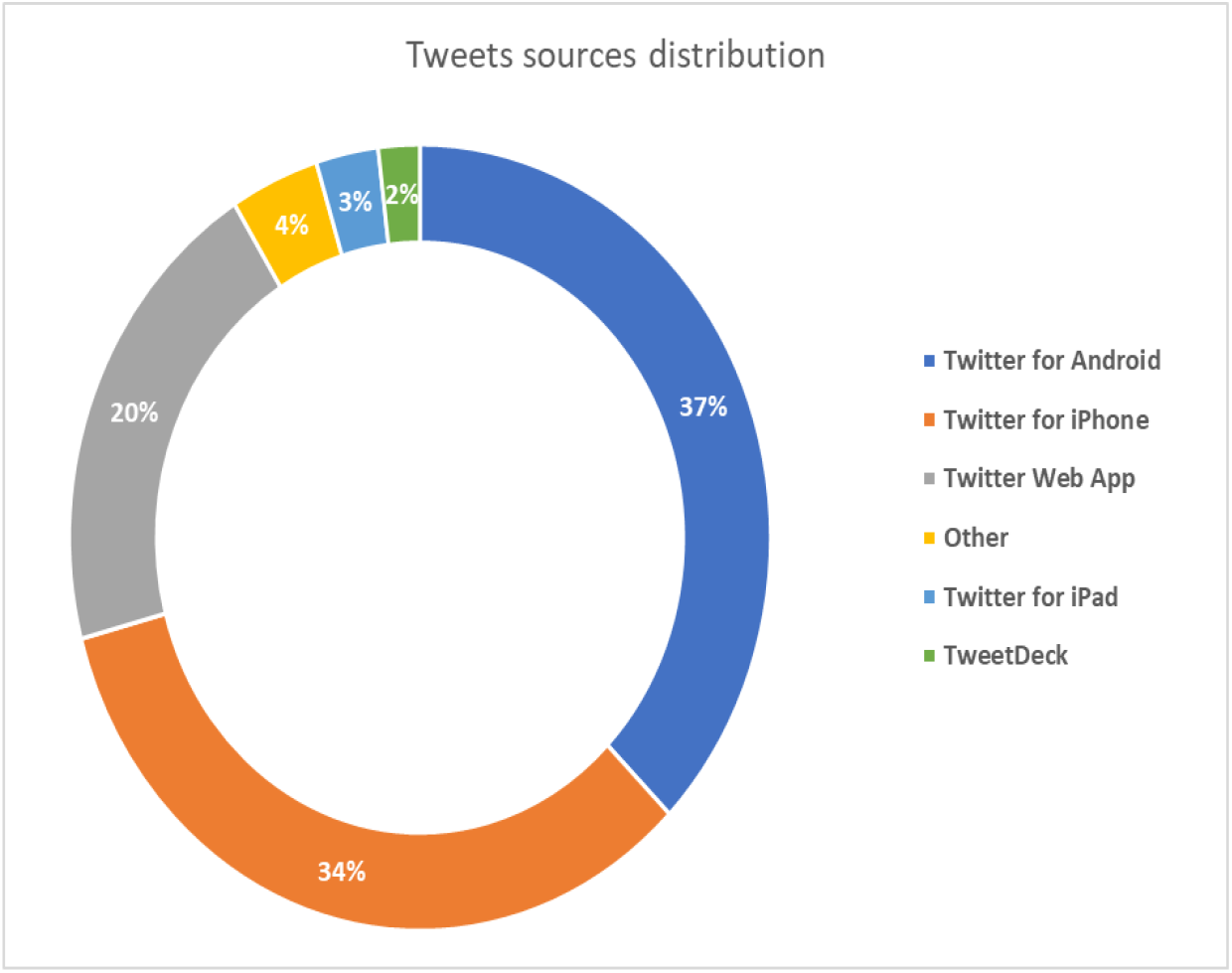
The tweets sources.

### 4.3. Top Keywords

The 25 top words are shown in Figure 3. These terms are eloquent as they are similar to the words expressed by the majority of people in real life. We observe four sets of words with approximatively the same respective frequency, they are cited in a decreasing order of their occurrence:

- virus, people
- Flu, Cases, Trump, test
- Spread, Health
- Work, outbreak, death, update, home, China, hands, report, world, country, call, confirmed, infected, week, risk, panic, pandemic

We can remark that these words describe all the situation people are overcoming in real life during this hard period and that we can summarize in a few paragraphs.

**Figure 3.**
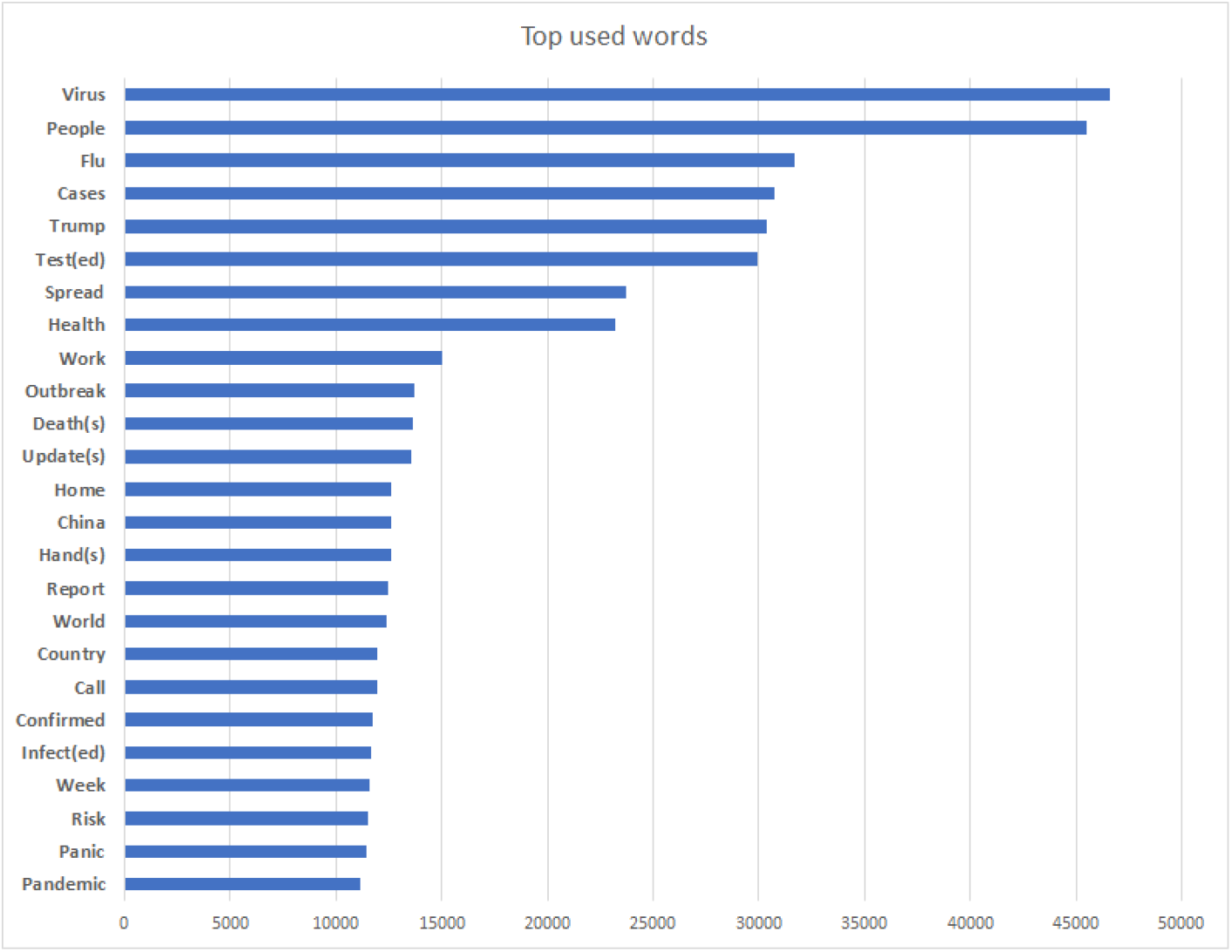
The 25 Top words of the dataset.

### 4.4 Tweets number per date

The number of tweets by date experienced the results shown in Figure 4. Regarding the number of published tweets, we find it significant during the period of this study and is changing from day to day. We remark that before the advent of this virus in December 2020 in China, very few tweets on coronavirus were published. These tweets consist in old tweets from the previous coronavirus versions that struck Asia in 2013, that were retweeted again by some users. Figure 5 depicts the boxplot of the data collected during the study period, where we see a non-asymmetric distribution of the tweets number and six outliers corresponding the following days: 03/01/2020, 03/12/2020, 03/15/2020 for the lowest numbers of tweets and 03/06/2020, 03/14/2020 and 03/21/2020 for the highest numbers of tweets.

**Figure 4.**
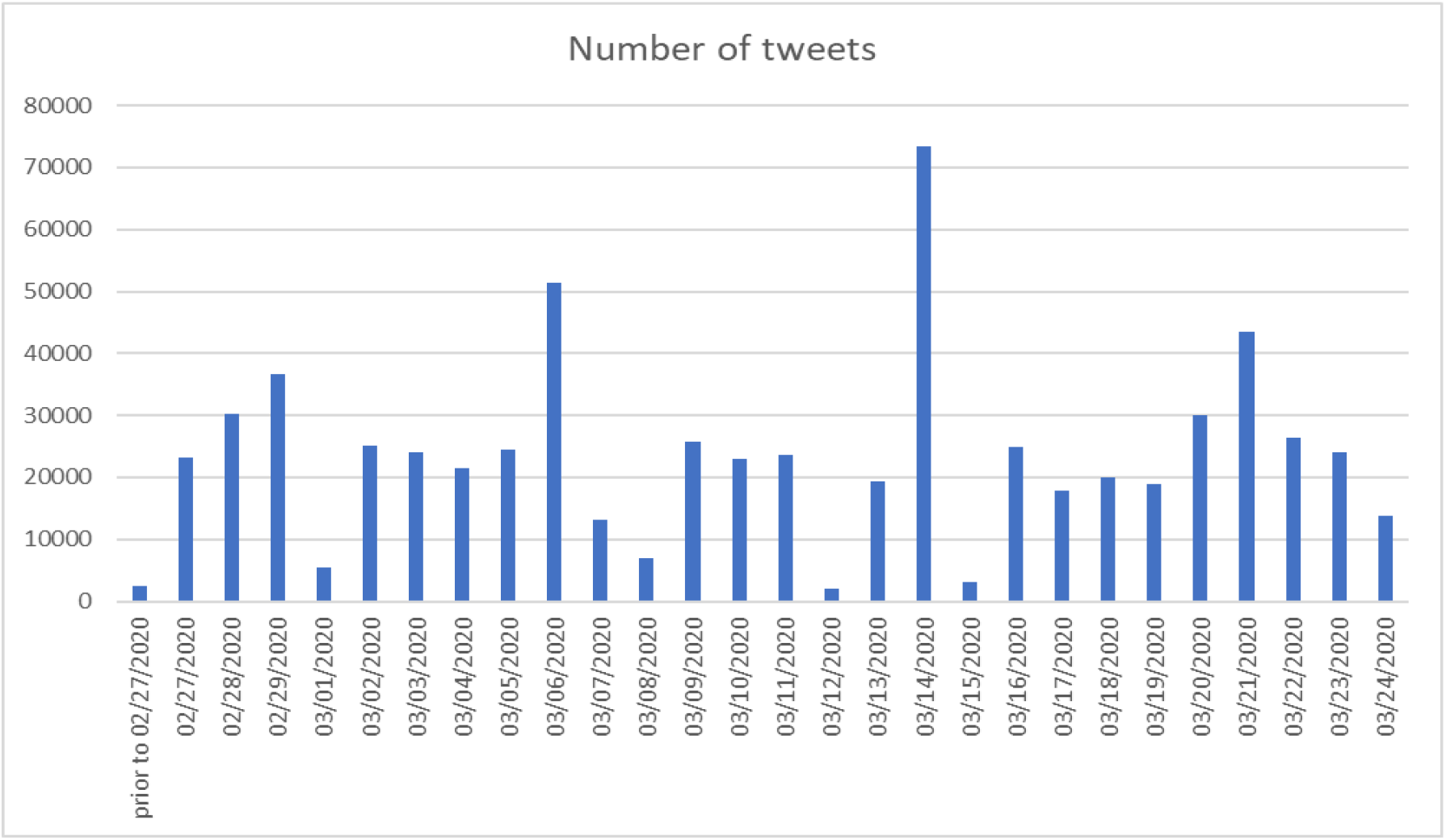
Number of tweets by date.

**Figure 5.**
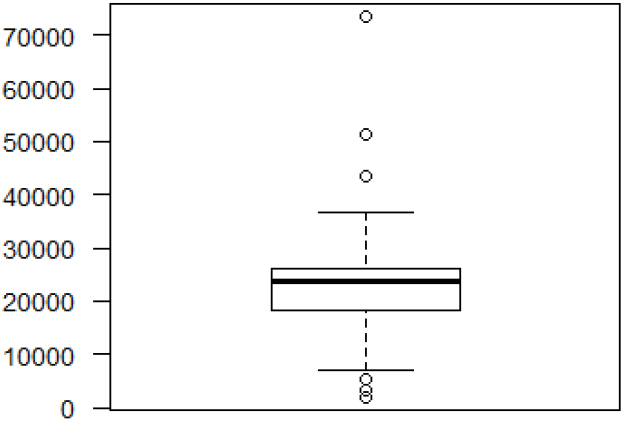
Boxplot on the number of tweets per day.

### 4.5. Number of Tweets per country

**The first ascertainment about the origin of the tweets is that a significant number of tweeters do not indicate their country. Therefore, the results hereafter are valid only for those tweets with a provenance information. Figure 6 shows the number of tweets per country where we observe that the majority of the published tweets emanate from USA. The second remark is that tweeters belong to several countries from all the continents. Note that the number of tweets is for some countries as important as their respective demography. For instance, India is ranked 3th for the number of tweets while it has the second largest population in the world. The same observation can be done for Nigeria and South Africa that count respectively the largest population in Africa. Also, the countries the most affected by the virus such as Spain, France and Italy are present in this graph**.

**Figure 6.**
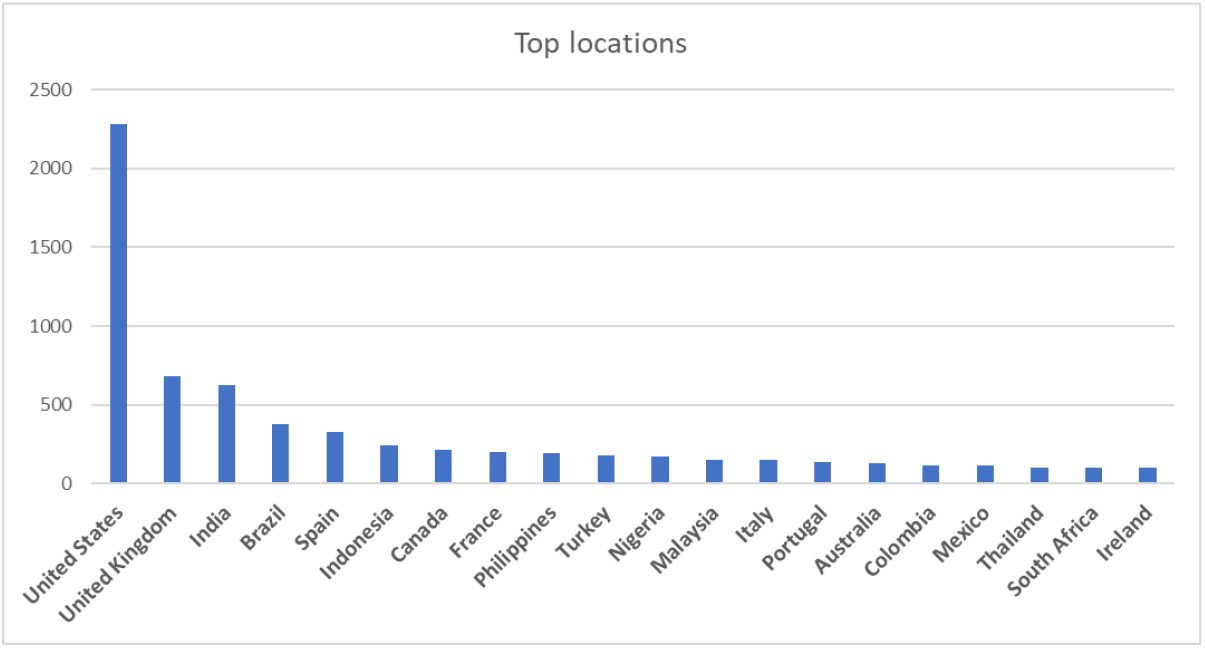
Number of tweets by country.

### 4.6. Tweets Language

The language of tweets has also been explored, Figure 7 gives an insight about this feature. We note that English prevails over the other languages. This can be explained by the fact that according to the previous statistics, the three highest amount of tweets are originated from the USA, United Kingdom and India, where the communication language is English. Also note that English is scientifically and technologically universal and is written by a large population of other countries. Spanish is ranked 2 in this graph because it is spoken in several countries like Colombia and Mexico that appear in the previous figure. Also the languages of the affected countries with a high degree during this period except China exist all in this graph.

**Figure 7.**
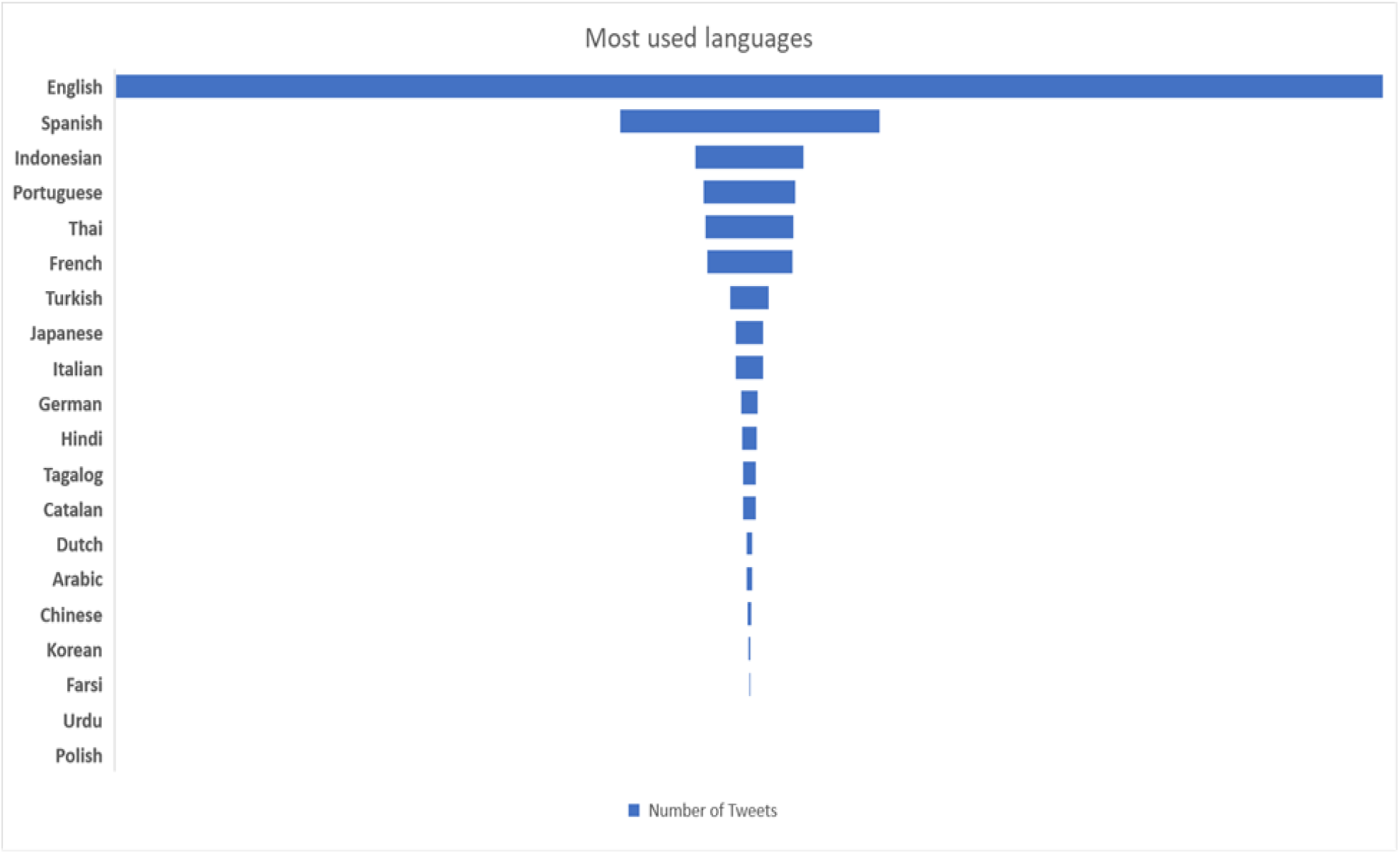
Number of tweets per language.

### 4.7 Correlations

We can think about several possible correlations between the parameters identified previously. In this subsection, we investigate the correlation between the number of real infected cases and the number of tweets published each day. The intuition is that the evolution of the number of infected cases urges people to communicate via the social media. Figure 10 depicts the case of the UK, where in the x axis, we have the number of tweets per day and in the y axis the number of the new cases tested positive to the coronavirus per day. We observe a positive trend for the correlation confirmed by a Pearson coefficient equal to 0,55.

**Figure 10.**
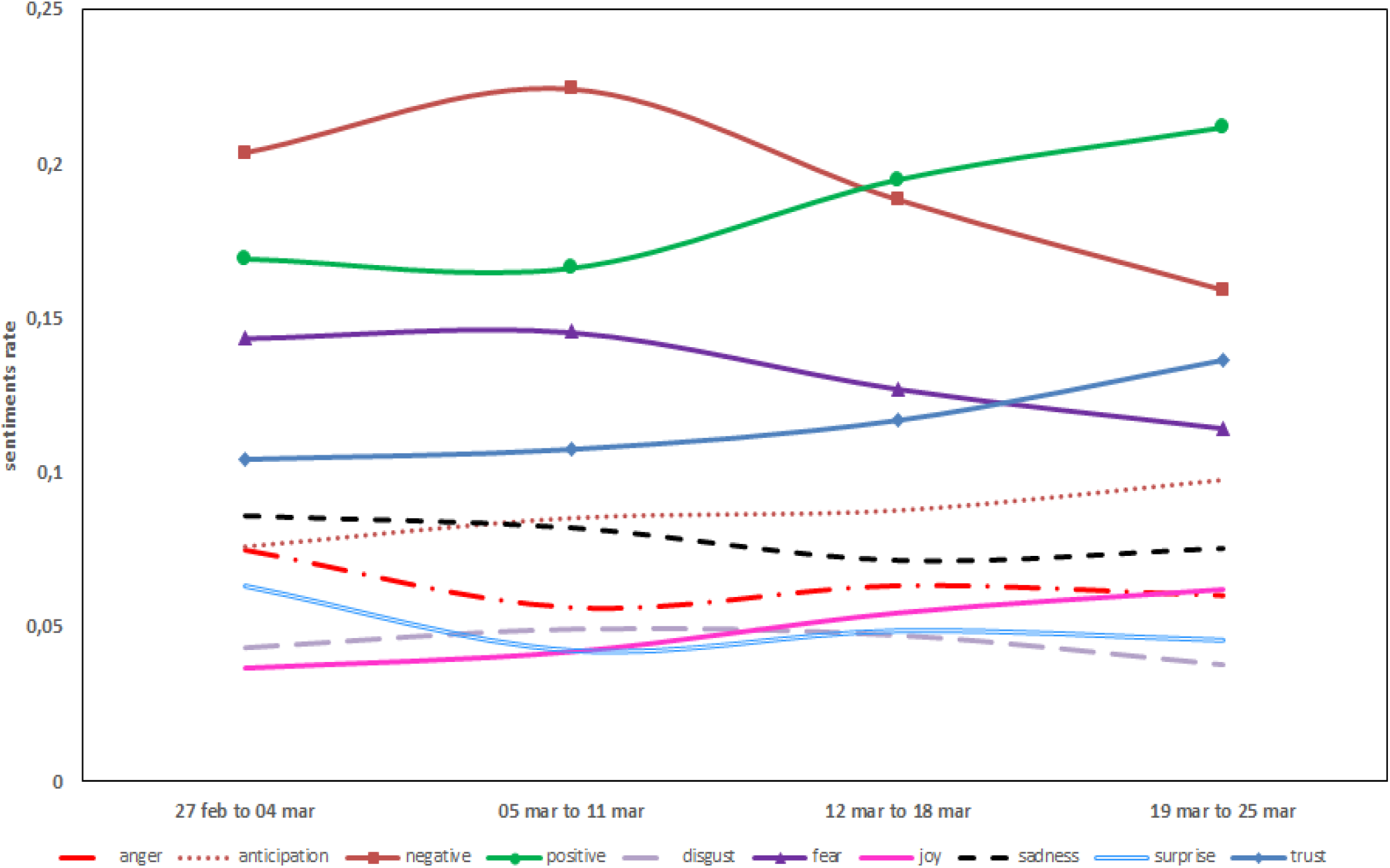
Correlation between the number of new cases and the number of tweets in the UK.

## 5. Tweets Sentiments Analysis

In addition to the previous study, a sentiment analysis was investigated aiming at capturing the tone of the tweets. We know that during the COVID-19 period, people changed their behaviors and experienced different feelings and emotions than those they have known before. Their lifestyle has shifted to something new. The algorithm described in the next subsection was adopted and implemented to shed insight on tweeters sentiments.

### 5.1. Sentiment Analysis Algorithm

The sentiment analysis algorithm we propose is based on the lexicon-based approach. As input, it takes the set of the preprocessed words of the dataset and a sentiment lexicon. There are several known sentiment lexicons that can be used for sentiment analysis. As examples we cite AFINN, Bing and NRC. AFINN computes a score from the range [-5,5] to each word and deducts the positive sentiment if the score is positive and the negative sentiment otherwise. Bing assigns straightly a positive or a negative sentiment to each word. NRC considers ten categories of sentiments, which are positive, negative, anger, anticipation, disgust, fear, joy, sadness, surprise, and trust and assigns to each word at least one of these categories. We chose the NRC lexicon as it provides specific sentiments other than positive and negative. This lexicon contains 6468 words distributed over the ten categories. The number of words in each category is shown in Table 2.

**Table 2.** Categories and their respective cardinalities in NRC.

| #words | Anger | Anticipation | Disgust | Fear | Joy | Negative | Positive | Sadness | Surprise | Trust |
| --- | --- | --- | --- | --- | --- | --- | --- | --- | --- | --- |
| 6468 | 1247 | 839 | 1058 | 1476 | 689 | 3324 | 2312 | 1191 | 534 | 1231 |

The framework of the algorithm is shown in Figure 8. After the preprocessing step, the list of the words of the whole dataset is determined. Meanwhile, the inverted file of the NRC is built and converted to a hash table. Initially each category is affected a count set to 0. Then the process considers the words one by one and calculates its hash key to access to the index that contains the address of the word in the lexicon. Once identifying the entry of the lexicon inverted file containing the word, it captures the categories and increments the count of each of them. Once the process is completed, the score for each category is computed as the ratio of its corresponding count over the total number of words.

Figure 9 shows the results achieved by the execution of the algorithm, that illustrate the tone of the tweets captured by the sentiment analysis. We observe that the negative sentiment has a score slightly greater than that of the positive. Fear has an important score whereas joy and disgust have the lowest. Although this sad situation, the tweeters seem to have trust in gaining the battle against the virus.

**Figure 8.**
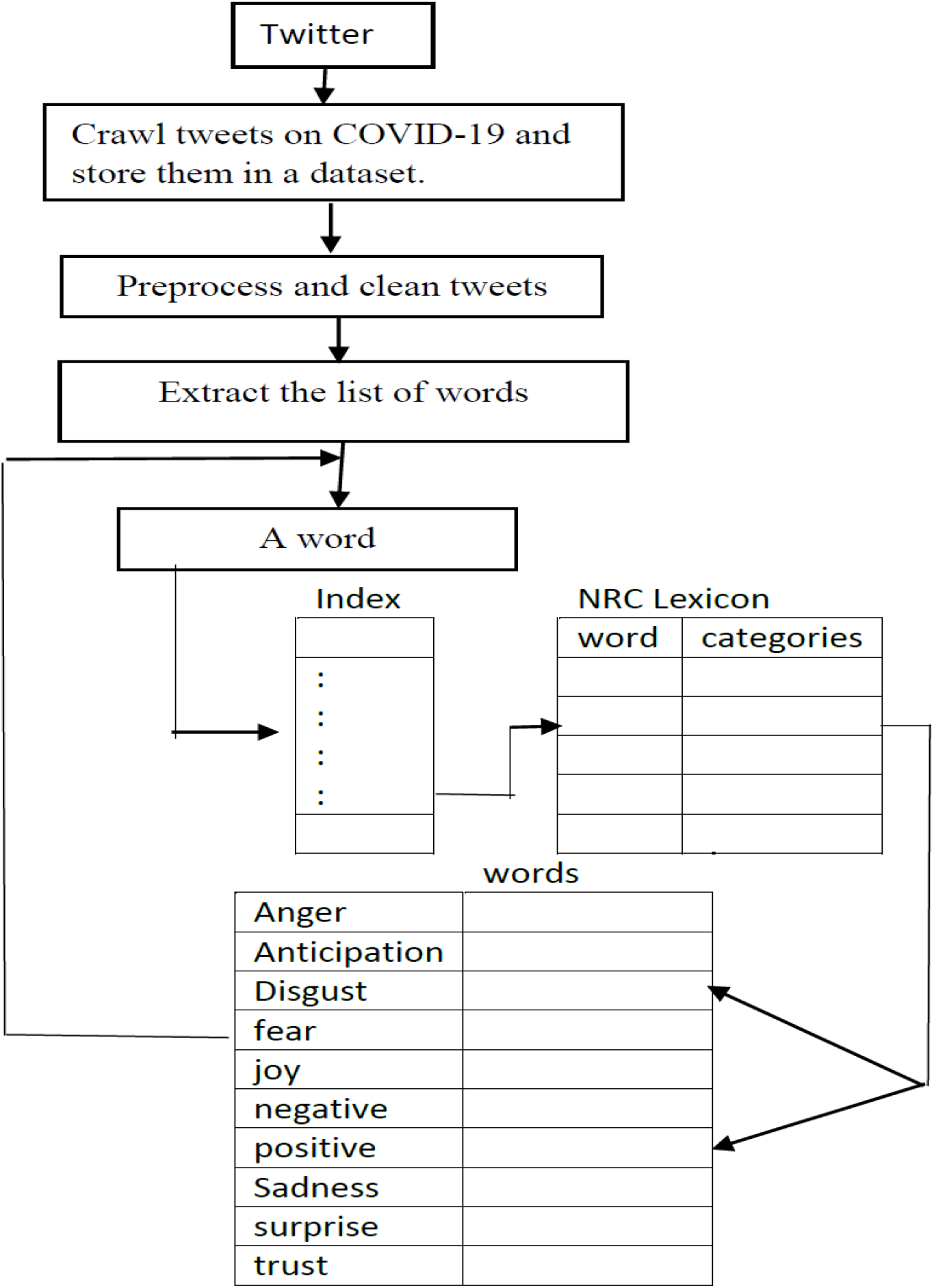
The flowchart of the sentiment analysis algorithm.

**Figure 9.**
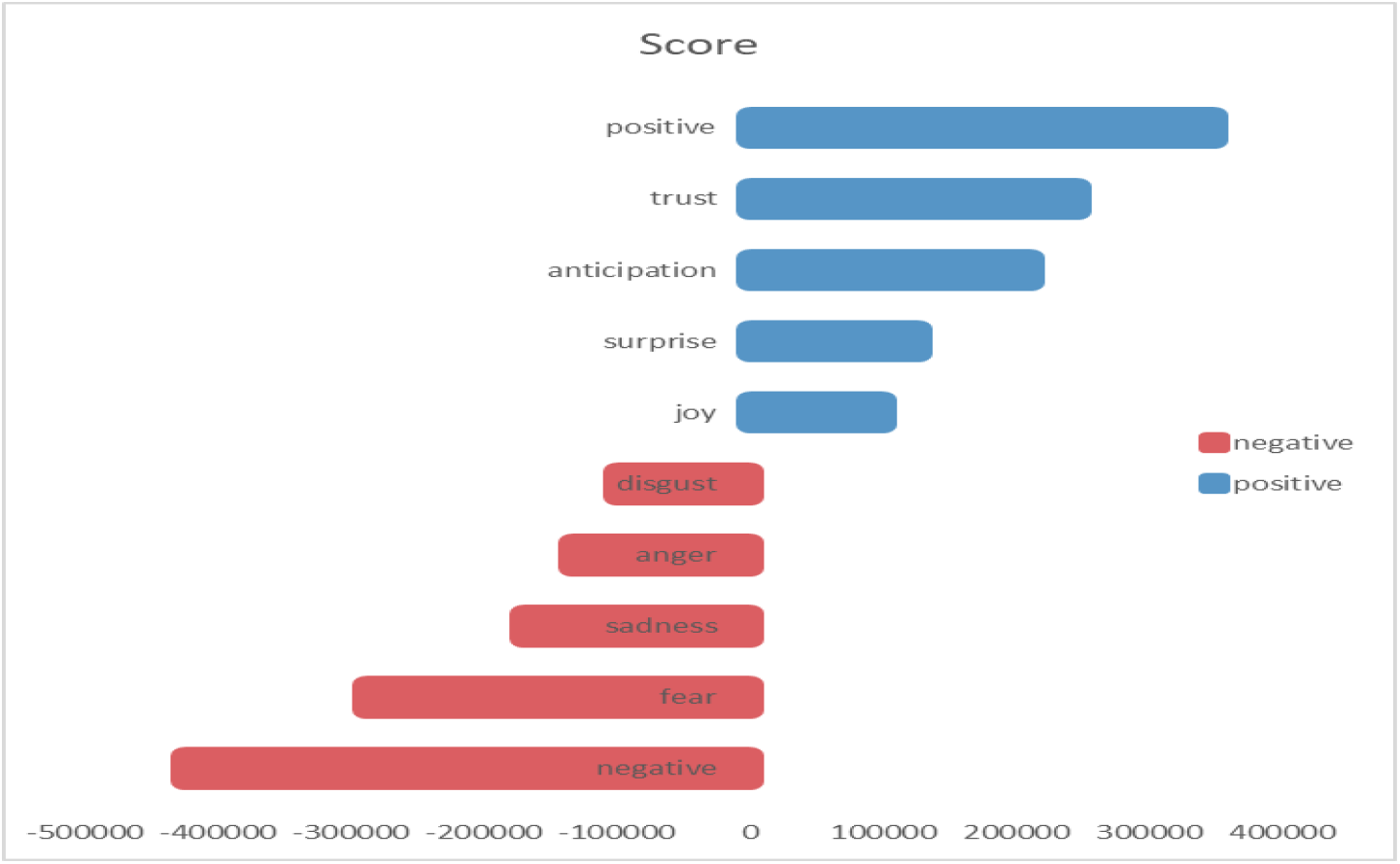
The tone of tweets captured by the sentiment analysis.

### 5.2. Sentiments Evolution

Figure 10 details the evolution of the sentiments during the four weeks. We notice that the *negative* sentiment was high in the beginning of the period then it knew a little increase in the second week and since then it is decreasing in an important degree. On the contrary, the *positive* sentiment follows the opposite behavior. *Fear* has almost the same evolution as the *negative* sentiment but with less magnitude whereas *trust* behaves as the *positive* sentiment with less importance. *Joy* is constant and is at the lowest score, which translates what people are feeling during these days.

**Figure 10.**
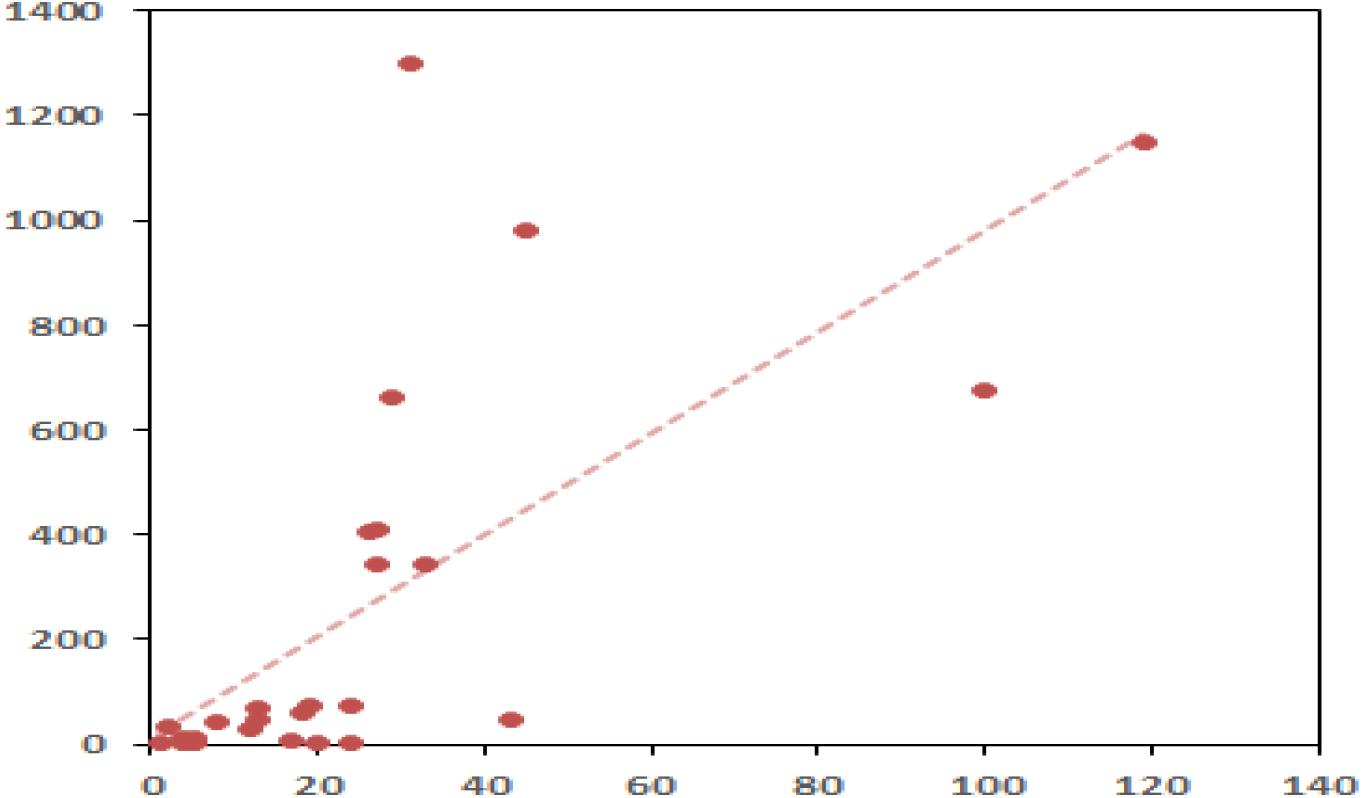
Sentiments Evolution during the four weeks.

### 5.3. Sentiments Analysis by country

As tweets can be distributed by country as shown in Figure 6, the sentiments analysis can be carried out for each country and each region by merging the tweets of the countries of the region in one dataset. Examples for such task were performed for the USA and Canada separately and the region of North America. India has also undergone such treatment. Note that the sentiments evolution can also be drawn for countries and regions.

## 5. Tweets Frequent Patterns and Association Rules Mining

Data mining technology proposes a large spectrum of tools to extract interesting and potentially useful patterns from huge and complex volumes of data. Association Rules Mining (ARM) is one of those techniques that can be adapted to various domains such as health, commerce and industry. The generation of ARM is achieved within two steps:

- Discovering Frequent Patterns or itemsets relative to a minimum support.
- Filtering ARM from the extracted frequent patterns with respect to a minimum confidence

The patterns for the tweets correspond to the most frequent words used in the tweets and hashtags, which represent all insights on this social media for the COVID-19.

Since the algorithms for Frequent Pattern Mining (FPM) are computationally expensive, a lot of research has been carried on further improving their effectiveness and efficiency. And especially, in order to cope with the main drawbacks of the Apriori and Eclat algorithms, the FP-Growth algorithm was developed [6]. The latter was then selected to be adapted to our tweets dataset.

### 5.1 FP-Growth for generating frequent tweets patterns

The FP-growth algorithm draws its strength from the fact it uses a sophisticated and optimal data structure called FP-tree that condenses only relevant data in a vertical format [6]. By scanning the tree, the frequent words respecting the minimum support are determined. The algorithm for mining frequent tweets keywords is outlined in Figure 11.

**Figure 11.**
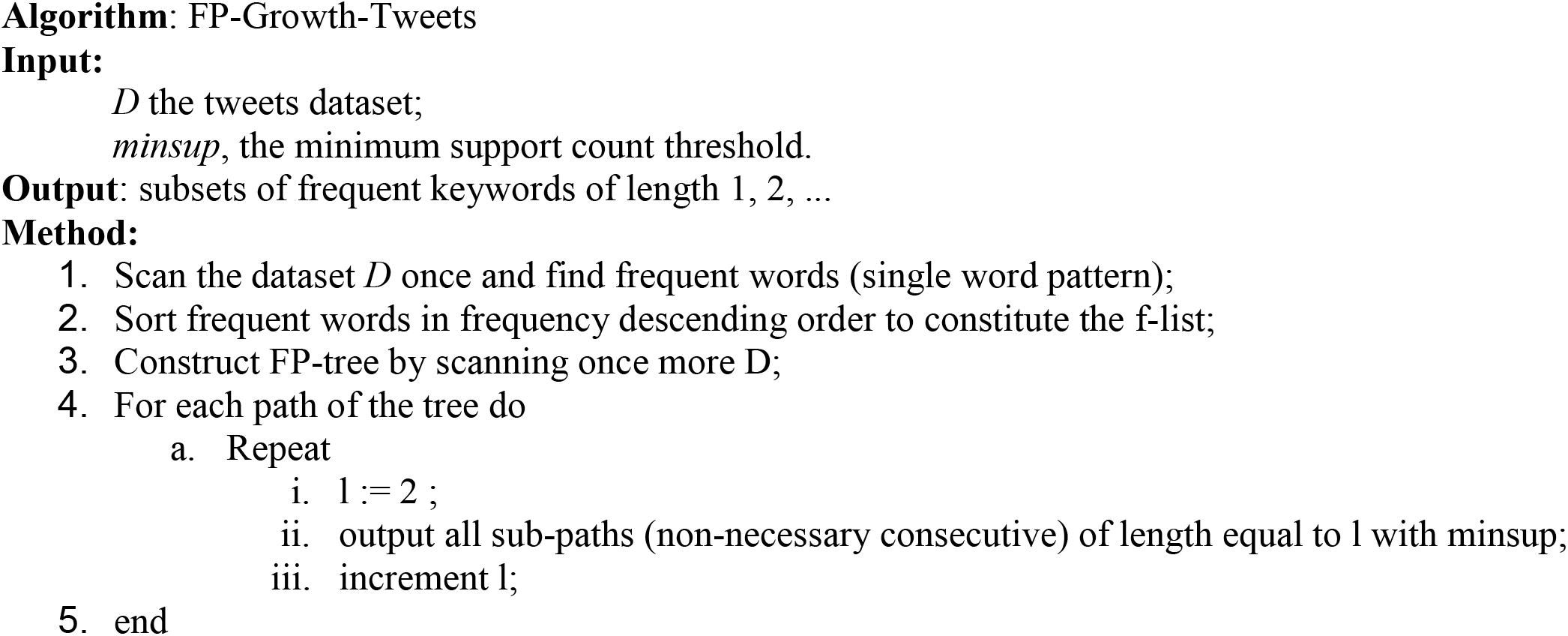
FP-Growth to mine twitter Data on COVID-19.

The advantage of the f-list is to eliminate at the beginning those words that do not respect the minimum support. Then the construction of the FP-tree considers only the *f-list* words and is consequently a data structure reduced to only relevant information. It starts with an empty node, then it creates nodes taken sequentially from the *f-list* and linking them if they belong to the same tweet while scanning the collection of tweets and incrementing their frequency.

### 5.2 Association Rules Mining for Tweets on COVID-19

Association rules are derived from the frequent patterns calculated by the adapted FP-Growth algorithm. Suppose {w1, w2, w3} are words appearing frequently in the tweets dataset with a minimum support equal to *minsup*. Then from this subset of words (called itemset in the traditional frequent patterns mining algorithms), we can generate 4 association rules that are:

→ w_1_, w_2_, w_3_ *(minsup, minconf)* w_1_ → w_2_, w_3_ *(minsup, minconf)* w_1_, w_2_, →w_3_ *(minsup, minconf)* w_1_, w_2_, w_3_ → *(minsup, minconf)*

The first rule means that in all tweets, the three words exist. In the second one, the rule is interpreted as: whenever w_1_ exists in a tweet, w_2_ and w_3_ appears in the same tweet. The other rules share the same meaning.

Note that we need to specify another measure for the rule besides the support, which is the confidence. The confidence of a rule is defined to be the probability whenever an antecedent of the rule is in a tweet, the consequent is also in the same tweet. It is calculated as:

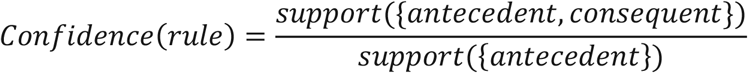

### 5.3. Experimental Results

The adapted FP-growth algorithm was implemented in Java programming language under the following environment: Windows 10 Intel core 15-7300U CPU at 2.60 Ghz, 8GB of RAM.

The most frequent words in the tweets published in English and calculated by the algorithm are reported in Table 3 in decreasing order of their support. We can confirm that these words constitute an important part of the daily vocabulary used by people nowadays in real life.

**Table 3.** The most frequent patterns with their minimum support.

| Item | MinSup |
| --- | --- |
| people | 26214.0 |
| virus | 22830.0 |
| cases | 19118.0 |
| flu | 18363.0 |
| tests | 17163.0 |
| spread | 14650.0 |
| health | 13574.0 |
| trump | 11149.0 |
| time | 10104.0 |
| work | 9627.0 |
| covid | 9511.0 |
| make | 8731.0 |
| outbreak | 8629.0 |
| pandemic | 8319.0 |
| day | 8289.0 |
| deaths | 8265.0 |
| china | 8103.0 |
| prepar | 8030.0 |
| updat | 7954.0 |

Similarly, the pair of words the most frequent appear in Table 4, which shows at the same time examples of association rules ordered with respect to the minimum support on the total number of patterns equal to 19560. These rules are impressively similar to those of the daily life during the pandemic. For instance, the rule *wash hand* has the highest confidence, which corresponds exactly to what we hear many times a day.

**Table 4.**
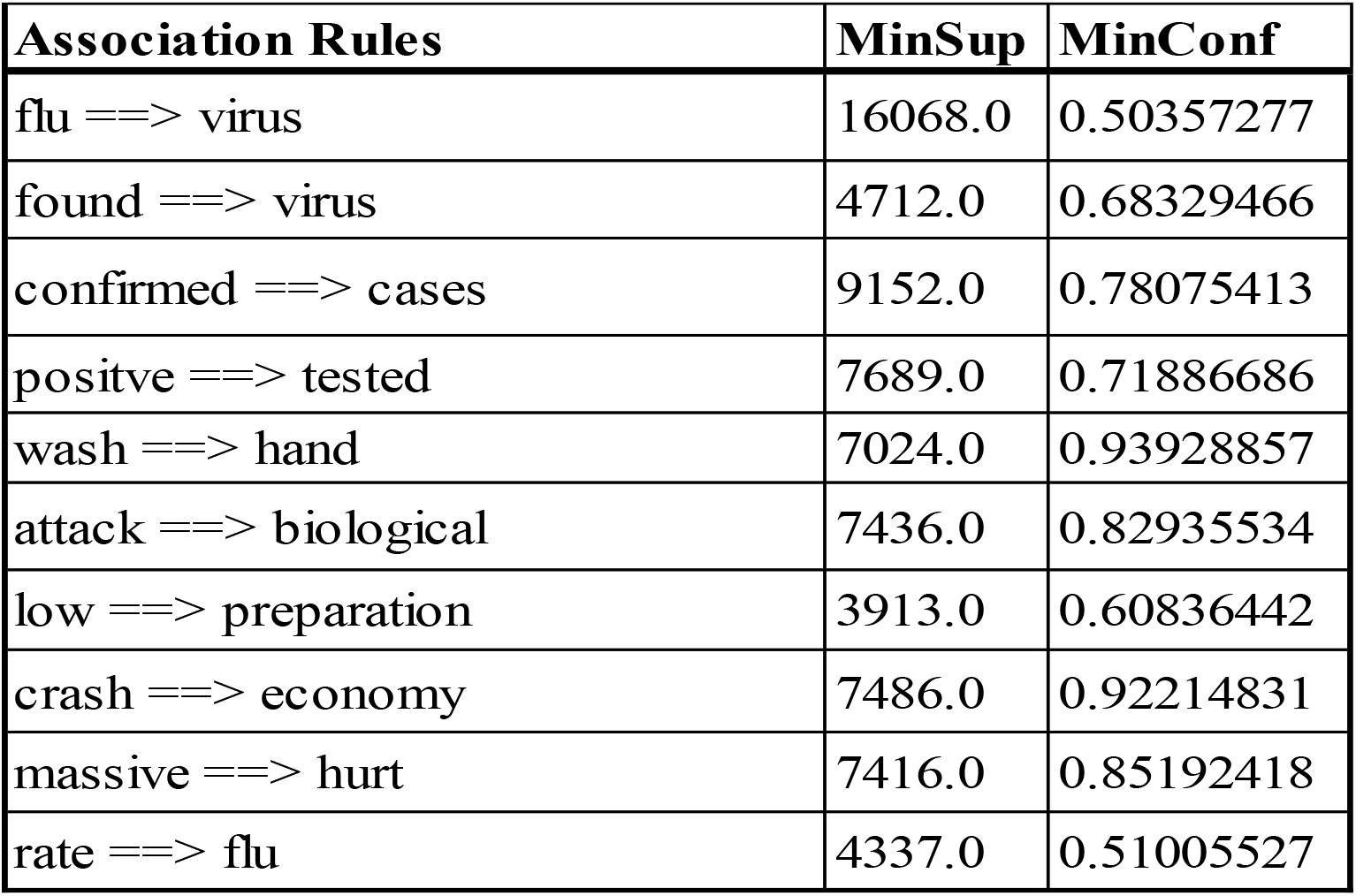
Examples of generated association rules with 2 frequent patterns.

Table 5 and Table 6 show the association rules identified by the algorithm for respectively the 3 and 4 most frequent patterns with a low support but a significant confidence. These rules are also eloquent relatively to what people are living these days.

**Table 5.**
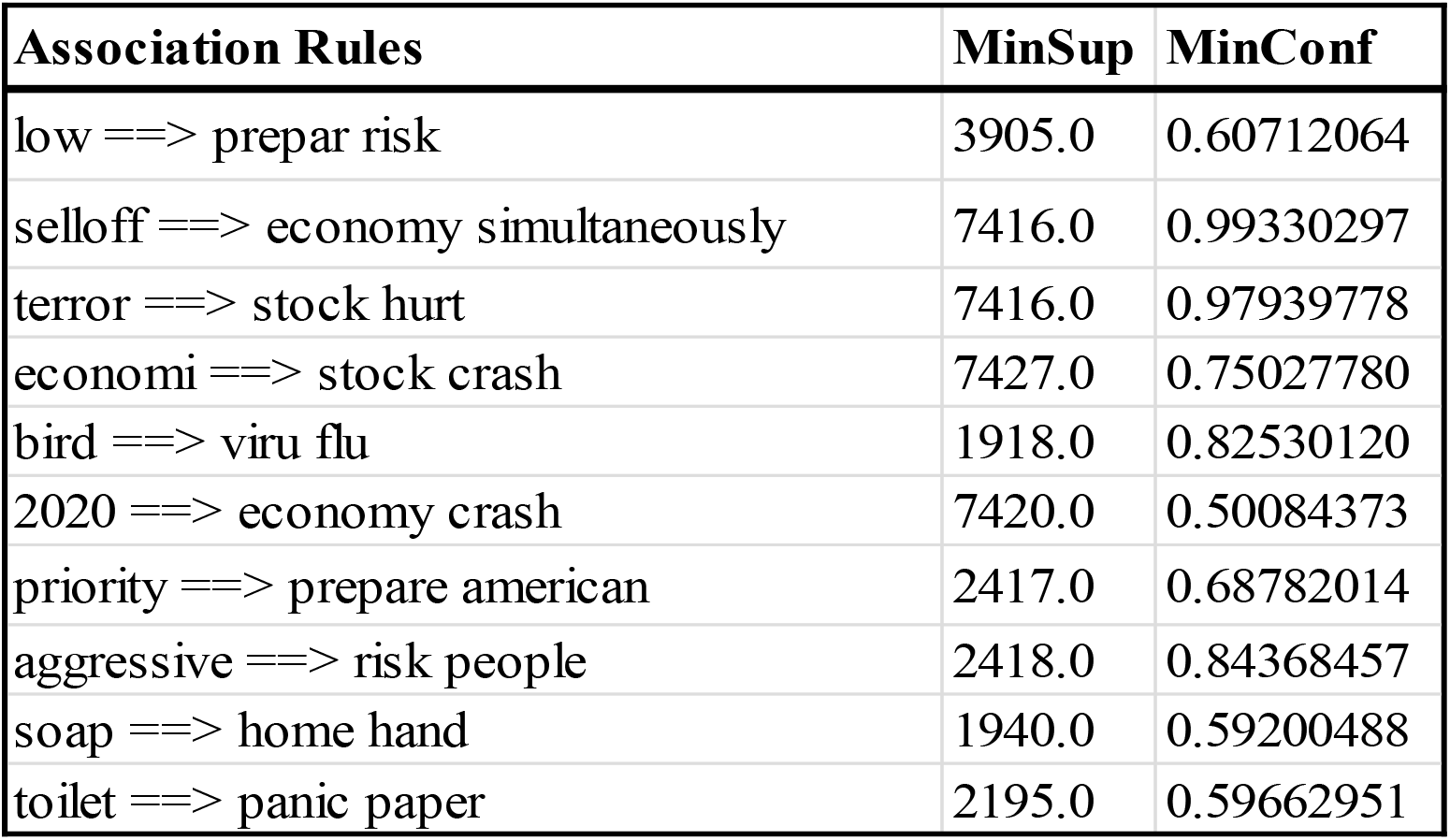
Examples of generated association rules with 3 frequent patterns.

| Association Rules | MinSup | MinConf |
| --- | --- | --- |
| low ==> prepar risk | 3905.0 | 0.60712064 |
| selloff ==> economy simultaneously | 7416.0 | 0.99330297 |
| terror ==> stock hurt | 7416.0 | 0.97939778 |
| economi ==> stock crash | 7427.0 | 0.75027780 |
| bird ==> viru flu | 1918.0 | 0.82530120 |
| 2020 ==> economy crash | 7420.0 | 0.50084373 |
| priority ==> prepare american | 2417.0 | 0.68782014 |
| aggressive ==> risk people | 2418.0 | 0.84368457 |
| soap ==> home hand | 1940.0 | 0.59200488 |
| toilet ==> panic paper | 2195.0 | 0.59662951 |

**Table 6.** Examples of generated association rules with 4 frequent patterns.

| Association Rules | MinSup | MinConf |
| --- | --- | --- |
| low ==> risk made american | 2414.0 | 0.41286129 |
| minister ==> prepare year india | 1480.0 | 0.42528735 |
| season ==> health doctor role | 1480.0 | 0.41398601 |
| men ==> season journey role | 1480.0 | 0.7834833 |
| secretary ==> update confirmed units | 1463.0 | 0.99659400 |
| communicating ==> update confirmed statesamerican | 1463.0 | 0.28924476 |
| communicating ==> updates confirmed families | 1463.0 | 0.2892447 |
| update ==> cases units low | 1463.0 | 0.18393261 |

## Conclusion and perspectives

Within the framework of this work, we were interested in investigating the people communication via social media during the terrible starting period of the rapid spread of COVID-19 all along the world. Twitter was selected to undertake the study because of the credibility of its users and also the ease of extracting the tweets. As a first contribution, a dataset of 653 996 tweets was extracted using *NodeXL* and preprocessed in order to highlight useful insights. As a second contribution, an exploratory study on the collection of tweets was carried out to yield descriptive statistics. A sentiment analysis study followed and interesting results were discussed. Data Mining technologies were afterwards exploited and the FP-Growth algorithm was especially adapted to the tweets dataset in order to discover the most frequent patterns in an effective and efficient way. The derived association rules highlight our understanding on the tweeters communicating on the COVID-19. The global results were impressive and can enrich the information forecasted by the traditional media.

As a future work, other measures such as the lift and the interestingness will be considered for the experiments of the frequent patterns mining with the purpose of improving the outcomes. Another possible research direction is to explore the way and the acceleration the virus has been spread around the word, Density-based clustering can be developed for this purpose.

## Data Availability

A dataset of more than 600 000 tweets on COVID-19 was constructed and is available. We can transmit it at any time.

---

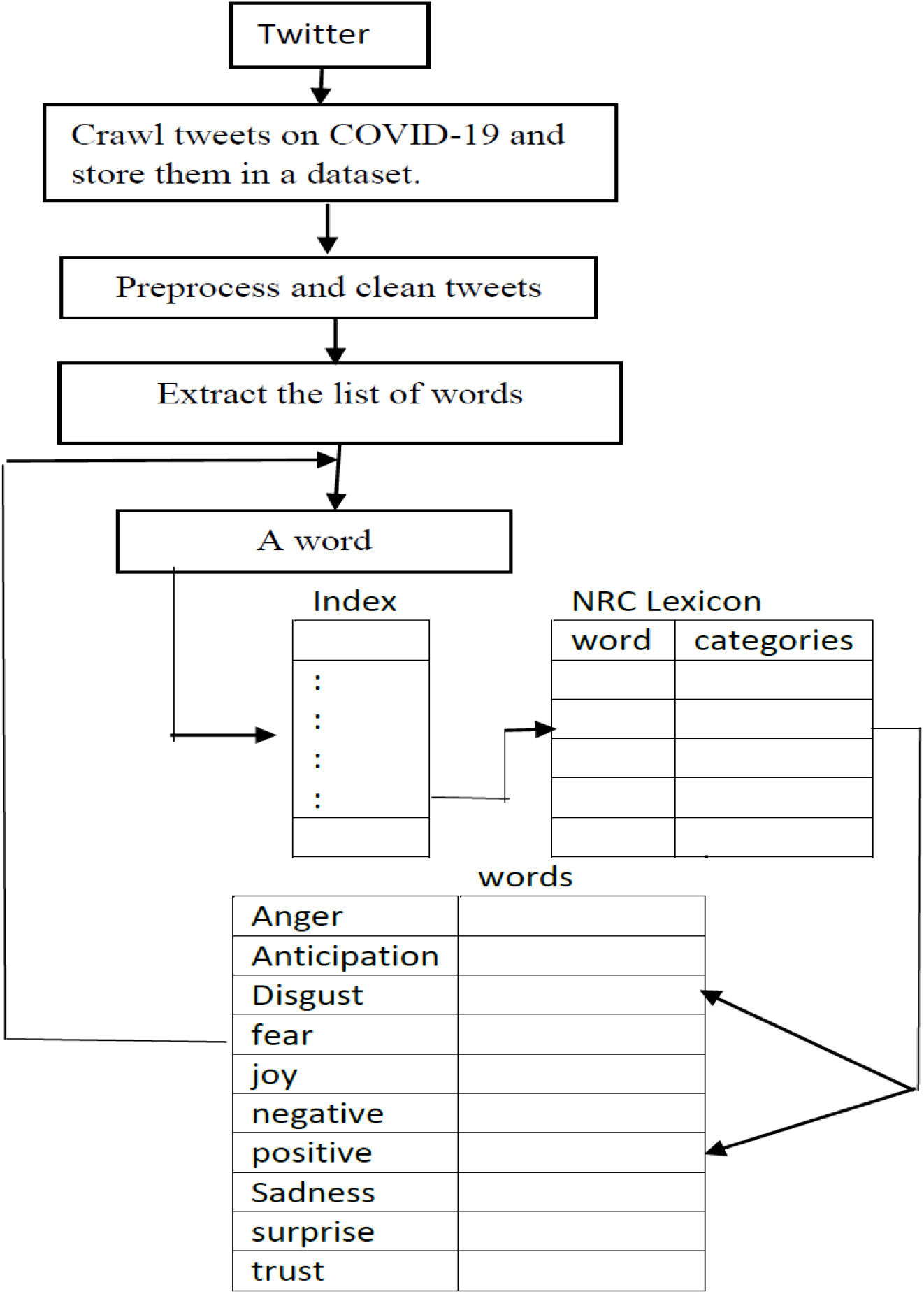

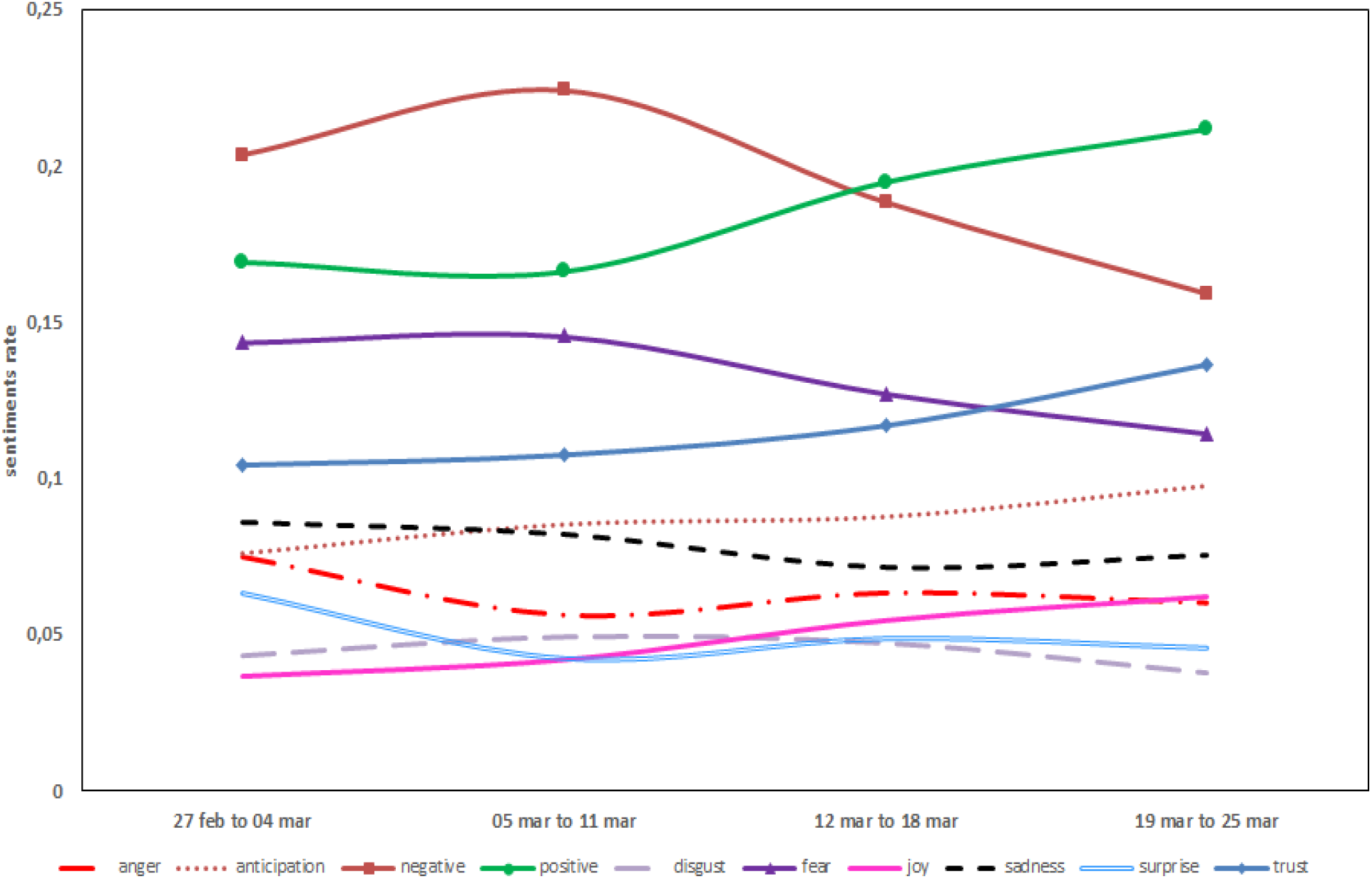

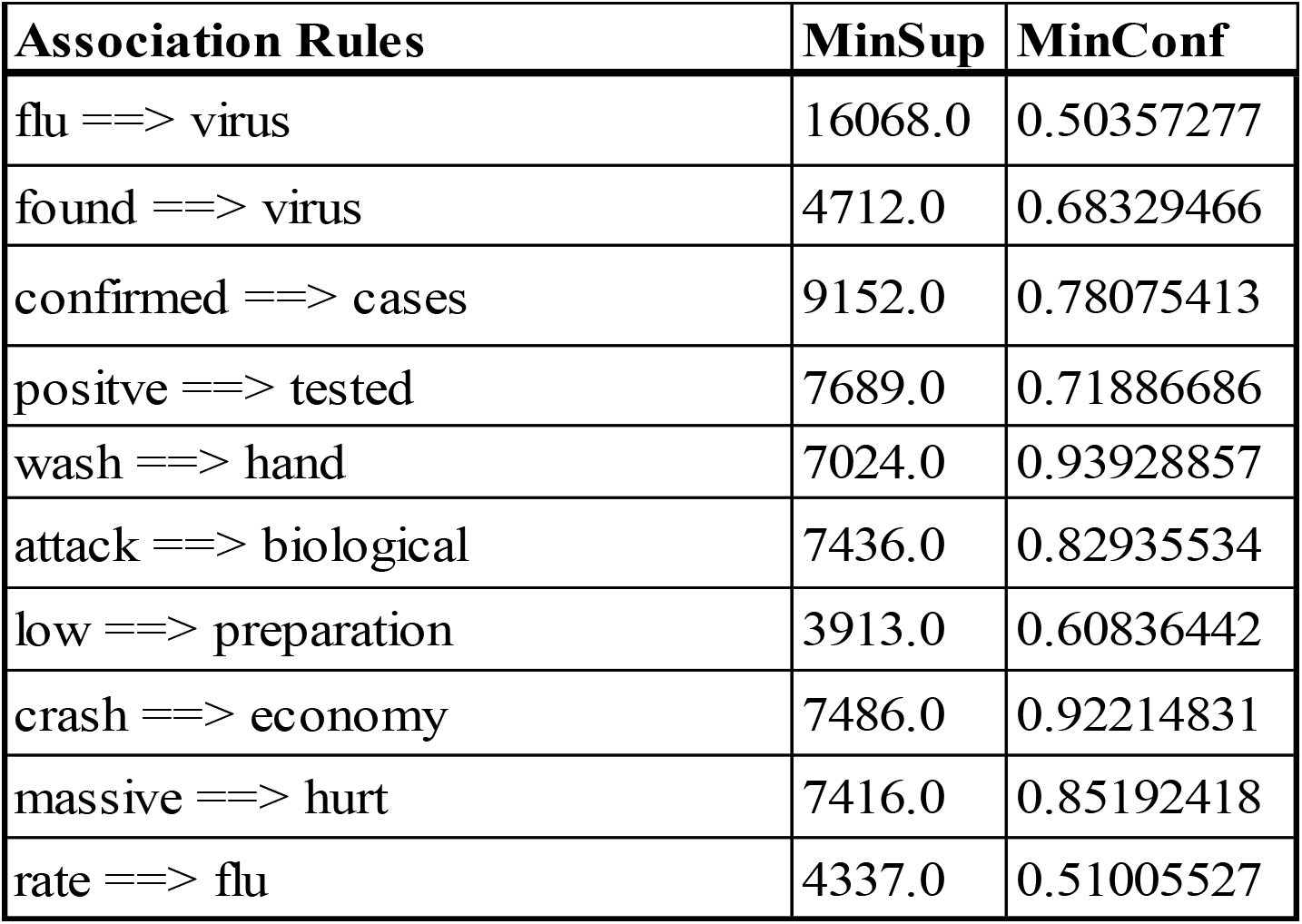

